# Prediction of American Society of Anesthesiologists Physical Status Classification from Preoperative Clinical Text Narratives Using Natural Language Processing

**DOI:** 10.1101/2023.02.03.23285402

**Authors:** Philip Chung, Christine T. Fong, Andrew M. Walters, Meliha Yetisgen, Vikas N. O’Reilly-Shah

## Abstract

**Importance:** Large volumes of unstructured text notes exist for patients in electronic health records (EHR) that describe their state of health. Natural language processing (NLP) can leverage this information for perioperative risk prediction.

**Objective:** Predict a modified American Society of Anesthesiologists Physical Status Classification (ASA-PS) score using preoperative note text, identify which model architecture and note sections are most useful, and interpret model predictions with Shapley values.

**Design:** Retrospective cohort analysis from an EHR.

**Setting:** Two-hospital integrated care system comprising a tertiary/quaternary academic medical center and a level 1 trauma center with a 5-state referral catchment area.

**Participants:** Patients undergoing procedures requiring anesthesia care spanning across all procedural specialties from January 1, 2016 to March 29, 2021 who were not assigned ASA VI and also had a preoperative evaluation note filed within 90 days prior to the procedure.

**Exposures:** Each procedural case paired with the most recent anesthesia preoperative evaluation note preceding the procedure.

**Main Outcomes and Measures:** Prediction of a modified ASA-PS from preoperative note text. We compared 4 different text classification models for 8 different input text snippets. Performance was compared using area under the receiver operating characteristic curve (AUROC) and area under the precision recall curve (AUPRC). Shapley values were used to explain model predictions.

**Results:** Final dataset includes 38566 patients undergoing 61503 procedures. Prevalence of ASA-PS was 8.81% for ASA I, 31.4% for ASA II, 43.25% for ASA III, and 16.54% for ASA IV-V. The best performing models were the BioClinicalBERT model on the truncated note task (macro-average AUROC 0.845) and the fastText model on the full note task (macro-average AUROC 0.865). Shapley values reveal human-interpretable model predictions.

**Conclusions and Relevance:** Text classification models can accurately predict a patient’s illness severity using only free-form text descriptions of patients without any manual data extraction. They can be an additional patient safety tool in the perioperative setting and reduce manual chart review for medical billing. Shapley feature attributions produce explanations that logically support model predictions and are understandable to clinicians.

## Introduction

Models to assess adverse event risk are indispensable tools in the arsenal of the perioperative clinician; guiding decision-making with respect to prehabilitation, preoperative testing, intraoperative management strategy, postoperative disposition, and more. These models base risk assessments on a limited number of discrete predictor variables. Examples include the American College of Surgeons (ACS) National Surgical Quality Improvement Program (NSQIP) Surgical Risk Calculator,^1,2^ the Revised Cardiac Risk Index (RCRI),^3,4^ and the Gupta Perioperative Risk for Myocardial Infarction or Cardiac Arrest (MICA).^5^ While well validated, classification of these predictor variables in many cases require expert clinician chart review and patient assessment. The use of these manually abstracted discrete data elements works well in the context of individual patient assessment. However, without *a priori* discretization of these elements, usage of these risk assessment tools for other purposes are limited. These purposes might include automation of risk assessment for patient safety purposes, perioperative population health assessment, benchmarking within and across health systems, or simply for triage of patients in assessing the need for a preoperative clinic visit.

Machine learning and natural language processing (NLP) techniques, coupled with adoption of electronic health records (EHR), and widespread availability of high-performance computational resources offer new avenues for perioperative risk stratification whereby unstructured data sources such as medical note free-form text may be directly input into prediction models without abstracting data elements. Unlike historical keyword-based approaches, modern NLP techniques using large pretrained language models are able to account for inter-word dependencies across the entire text sequence and have been shown to achieve state of the art performance on a variety of NLP tasks^6–9^ including text classification.^10,11^ However it is unknown whether these techniques can be successfully applied to perioperative risk prediction. In particular, we investigate risk prediction using only unstructured text notes written by clinicians drawn from the EHR, which often contain narratives that richly and concisely describe a nuanced clinical picture of the patient while simultaneously prioritizing the clinician’s pertinent concerns.

The American Society of Anesthesiologists Physical Status (ASA-PS) score ^12,13^ is a categorical clinician-driven assessment of patient periprocedural risk. ASA-PS has been shown to be an independent predictor of mortality and patient outcomes^14–19^ despite well-described interrater variability in ASA-PS classification.^20,21^ In this study, we investigated prediction of ASA-PS directly from free-form text taken from an anesthesia preoperative evaluation note using four different text classification approaches that span the spectrum of historical and modern techniques: (1) random forest^22^ with n-gram and term-frequency inverse document frequency (TFIDF) transform,^23^ (2) support vector machine^24^ with n-gram and TFIDF transform, (3) fastText^25,26^ word vector model, and (4) BioClinicalBERT deep neural network language model. We compared the model’s prediction with the ASA-PS assigned by the anesthesiologist on the day of surgery and hypothesized that advanced NLP modeling techniques would provide improved predictions of ASA-PS score as compared to simpler models.

## Methods

This retrospective study of routinely collected health records data was approved by the University of Washington Human Subjects Division with a waiver of consent. This study followed the Transparent Reporting of a Multivariable Prediction Model for Individual Prognosis or Diagnosis (TRIPOD) guideline^27^ and other guidelines specific to machine learning projects.^28–30^ eFigure 1 depicts a flow diagram of study design.

### Study Cohort

Inclusion criteria were patients who had a procedure requiring anesthesia at the University of Washington Medical Center or Harborview Medical Center from January 1, 2016 – March 29, 2021 where the patient also had an anesthesia preoperative evaluation note filed up to 6 hours after the anesthesia end time. This 6-hour grace period reflects the reality that in some urgent or emergency situations or due to EHR behavior, text documentation may be time stamped out of order.

The note must have contained the following sections: History of Present Illness (HPI), Past Medical and Surgical History (PMSH), Review of Systems (ROS), and Medications; notes missing at least one of these sections were excluded. Cases must have had a recorded value for ASA-PS assigned by the anesthesiologist of record, a free-form text Procedure description, and a free-form text Diagnosis description; cases missing at least one of these values are excluded.

A unit of analysis is defined as a single case with an anesthesia preoperative evaluation note filed within 90 days of the procedure. This unit was chosen because ASA-PS is typically recorded on a per-case basis by the anesthesiologist to reflect the patient’s pre-anesthesia medical comorbidities at the time of the procedure. Likewise, preoperative evaluation notes filed >90 days before the case are not considered to reflect the patient’s state of health so are excluded. Data was randomly split 70%-10%-20% into training, validation, and test datasets respectively. Patients with multiple cases were randomized into a single data split to avoid information leakage between the three datasets. New case number identifiers were generated for this study and used to refer to each case.

### Outcomes

The outcome variable is a modified ASA-PS with valid values of ASA I, ASA II, ASA III, ASA IV-V. ASA V cases are extremely rare, resulting in class imbalances that affect model training and performance. Thus ASA IV and V were combined into a compound class “IV-V”. ASA VI organ procurement cases are excluded. The final categories retain the spirit of the ASA-PS for perioperative risk stratification and resembles the original ASA-PS devised by Saklad in 1941.^12,31^ The emergency surgery modifier “E” was discarded.

### Predictors and Data Preparation

Free-form text from the anesthesia preoperative evaluation note is organized into many sections. Regular expressions are used to extract HPI, PMSH, ROS, and medications from the note. While diagnosis and procedure sections exist within the note, they were less frequently documented than in the procedural case booking data from the surgeon. Therefore, free-form text for these sections were taken from the case booking. Newline characters and whitespaces were removed from the text. Note section headers were excluded so that only the body of text from each section is included. We used text from each section to train models for ASA-PS prediction, resulting in 8 prediction tasks: Diagnosis, Procedure, HPI, PMSH, ROS, Medications (Meds), Note, Truncated Note (Note512). “Note” refers to using the whole note text as the predictor to train a model. When BioClinicalBERT is applied to the “Note” task, the WordPiece tokenizer^32–34^ truncates input text to 512 tokens. This truncation does not occur for other models. For equitable comparison across models, we define the “Note512” task, which truncates the note text to the first 512 tokens used by the BioClinicalBERT model.

### Statistical Analysis and Modeling

Four model architectures with different conceptual underpinnings were trained: (1) Random forest (RF),^22^ (2) Support vector machine (SVM),^24^ (3) fastText,^25,26^, and (4) BioClinicalBERT.^35^ Each model architecture was trained on each of the 8 prediction tasks for a total of 32 final models.

Each model was trained on the training dataset. Model hyperparameters were tuned using Tune^36^ with the BlendSearch^37,38^ algorithm to maximize Matthew’s Correlation Coefficient (MCC) computed on the validation dataset. The number of hyperparameter tuning trials was selected to be 20 times the number of model hyperparameters with early stopping if the MCC of the last 3 trials reaches a plateau with standard deviation <0.001. The best model was then evaluated on the held-out test dataset. Details on the approach taken for each of the four model architectures is available in supplemental methods.

### Baseline Models

Two baseline models were created for comparison: a random classifier model and an age classifier model. The random classifier model generates a random prediction without using any features, thus serving as a negative control baseline. The age classifier model is a simple multiclass logistic regression model with cross-entropy loss and L2 penalty that uses age to directly predict the modified ASA-PS outcome variable. Defaults were used for all other model parameters. Both baselines were implemented using Scikit-learn.

### Evaluation Metrics

Final models were evaluated on the held-out test dataset by computing both class-specific and class-aggregate performance metrics. Class-specific metrics include: receiver operator characteristic (ROC) curve, area under receiver operator curve (AUROC), precision-recall curve, area under precision-recall curve (AUPRC), precision (positive predictive value), recall (sensitivity), and F1. Class-aggregate performance metrics include MCC and AUCmu,^39^ a multiclass generalization of the binary AUROC. Additionally, macro-average AUROC, AUPRC, precision, recall and F1 were also computed.

### Model Interpretability and Error Analysis

4-by-4 contingency tables were generated to visualize the distribution of model errors. Catastrophic errors were defined as cases where the model predicts ASA IV-V but the anesthesiologist assigned ASA I, or vice versa. For catastrophic errors made by the BioClinicalBERT model with the Note512 task, three new anesthesiologist raters independently assigned an ASA-PS based on only the input text from the Note512 task. These new ASA-PS ratings were compared against the original anesthesiologist’s ASA-PS as well as the model prediction’s ASA-PS.

The SHAP^40^ python package was used to train a Shapley values feature attribution model on the test dataset to understand which words support prediction of each modified ASA-PS outcome variable. An analysis of model errors with Shapley value feature attributions was reviewed for each of the catastrophic error examples with representative examples included in the manuscript. Shapley values for predicting each ASA-PS are visualized as a heatmap over text examples. Text examples are de-identified by replacing ages, dates, names, locations, and entities with pseudonyms to achieve data obfuscation while preserving structural similarity to the original passage.

## Results

Our study comprised 38,566 patients undergoing 61,503 procedures with 46,275 notes. Baseline patient, procedure, and note characteristics are described in Table 1. A flow diagram describing dataset creation is shown in eFigure 2.

**Table 1:** Baseline patient, procedure, and note characteristics for Train, Validation, Test datasets.

|  |  |  | Train | Validation | Test |
| --- | --- | --- | --- | --- | --- |
| Patient Characteristics | Patient Count, no. (% across dataset splits) |  | 26994 (70.0%) | 3858 (10.0%) | 7714 (20.0%) |
|  | Number of Surgeries per Patient, no. (% within dataset split) | 1 | 19107 (70.78%) | 2741 (71.05%) | 5475 (70.97%) |
|  |  | 2 | 4528 (16.77%) | 608 (15.76%) | 1330 (17.24%) |
|  |  | 3 | 1635 (6.06%) | 249 (6.45%) | 425 (5.51%) |
|  |  | 4 | 715 (2.65%) | 124 (3.21%) | 224 (2.9%) |
|  |  | >=5 | 1009 (3.74%) | 136 (3.53%) | 260 (3.37%) |
|  | Age, mean (SD) |  | 50.59 (18.16) | 51.51 (18.09) | 50.66 (18.0) |
|  | Gender, no. (% within dataset split) | Female | 18419 (70.62%) | 2534 (9.72%) | 5130 (19.67%) |
|  |  | Male | 24720 (69.79%) | 3646 (10.29%) | 7053 (19.91%) |
|  |  | Unknown | 0 (0.0%) | 0 (0.0%) | 1 (100.0%) |
| Procedural Case Characteristics | Case Count, no. (% across dataset splits) |  | 43139 (70.14%) | 6180 (10.05%) | 12184 (19.81%) |
|  | Anesthesia Type, no. (% within dataset split) | General | 34901 (81.07%) | 4961 (80.51%) | 9927 (81.64%) |
|  |  | MAC | 7063 (16.41%) | 1005 (16.31%) | 1905 (15.67%) |
|  |  | Regional | 1089 (2.53%) | 196 (3.18%) | 327 (2.69%) |
|  | ASA Physical Status Classification Score, no. (% within dataset split) | I | 3734 (8.66%) | 555 (8.98%) | 1127 (9.25%) |
|  |  | II | 13631 (31.6%) | 1875 (30.34%) | 3806 (31.24%) |
|  |  | III | 18626 (43.18%) | 2649 (42.86%) | 5327 (43.72%) |
|  |  | IV-V | 7148 (16.57%) | 1101 (17.82%) | 1924 (15.79%) |
|  | Time Between Pre-Anesthesia Note and Surgery, median days HH:MM:SS (IQR) |  | 0 days 17:11:48<br>(0 days 00:17:00, 4 days 06:04:05) | 0 days 17:28:55<br>(0 days 00:18:00, 4 days 05:04:10) | 0 days 17:29:55<br>(0 days 00:17:05, 4 days 01:52:53) |
| Note Characteristics | Notes Count, no. (% across dataset splits) |  | 32444 (70.11%) | 4649 (10.05%) | 9182 (19.84%) |
|  | Text Word-Level Length, median (IQR) | Full Note | 727 (514, 999) | 723 (514, 1010) | 722 (511, 997) |
|  |  | Procedure | 5 (4, 8) | 5 (4, 8) | 5 (4, 8) |
|  |  | Diagnosis | 3 (2, 5) | 3 (2, 5) | 3 (2, 5) |
|  |  | HPI | 86 (35, 162) | 87 (35, 161) | 88 (35, 163) |
|  |  | PMSH | 28 (18, 42) | 28 (19, 44) | 28 (18, 42) |
|  |  | ROS | 87 (53, 154) | 87 (54, 155) | 87 (54, 153) |
|  |  | Medications | 145 (59, 264) | 143 (59, 264) | 146 (57, 262) |

AUROC for each model architecture and task is shown in Table 2; AUPRC is shown in eTable 1; AUC μ and MCC is shown in eTable 2. RF, SVM, and fastText perform best using the entire note compared to note sections. Tasks with longer text snippets yielded better performance–HPI, ROS and Meds sections result in better model performance as compared to Diagnosis, Procedure, and PMSH. On the Note task, fastText performs the best. On the Note512 task, BioCinicalBERT performs the best.

**Table 2:**
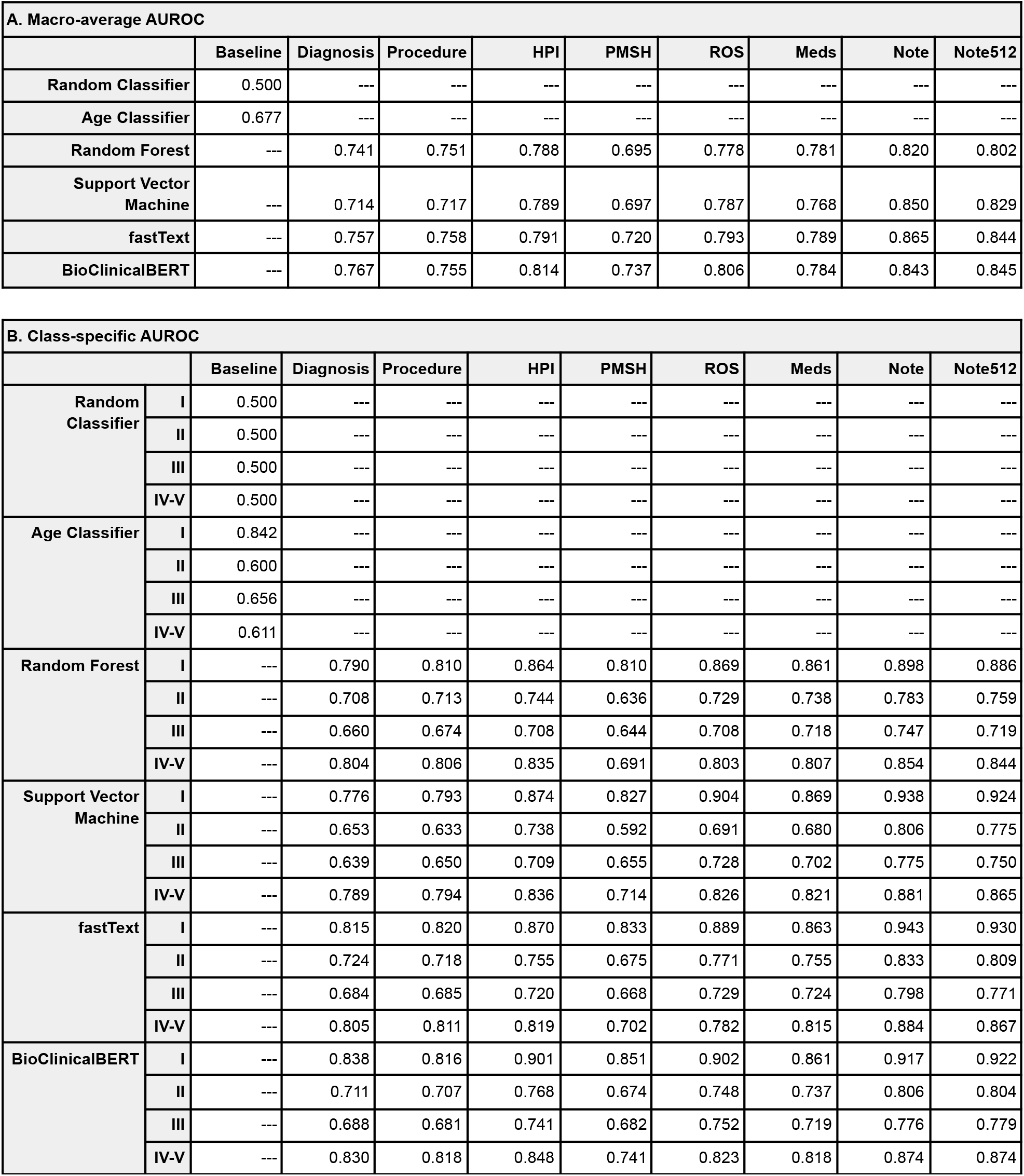
(A) Macro-average AUROC and (B) class-specific AUROC for each model architecture and task on the held-out test set compared to baseline models. Random Classifier serves as a negative control baseline. Age classifier serves as a simple clinical baseline since ASA-PS typically increases as a patient ages and has increased medical comorbidities.

Direct comparison of models is most appropriate using the Note512 task since all models are given the same information content. For this task, BioClinicalBERT has better class-aggregate performance across AUROC, AUPRC, AUCmu, MCC, F1 (eTable 3), recall (sensitivity) (eTable 5) metrics while the fastText model has better precision (positive predictive value) (eTable 4). Class-specific metrics also reflect this finding and show that fastText has high recall for ASA II and III, the most prevalent classes, but recall for ASA I and IV-V is considerably lower. BioClinicalBERT has similar or better AUROC and AUPRC across all the ASA-PS classes. This is also seen in the ROC curves (eFigure 4) and the precision-recall curves (eFigure 5), in which the BioClinicalBERT model generally shows better performance across most thresholds.

Figure 1 depicts 4-by-4 contingency tables to visualize distribution of model errors on the Note512 task. When erroneous predictions occur, they are typically adjacent to the ASA-PS assigned by the original anesthesiologist. For catastrophic errors made by the BioClinicalBERT model on the Note512 task, ASA-PS ratings from the three new anesthesiologist raters show greater concordance with the model’s predictions than the original anesthesiologist’s assignment (Figure 2).

**Figure 1:**
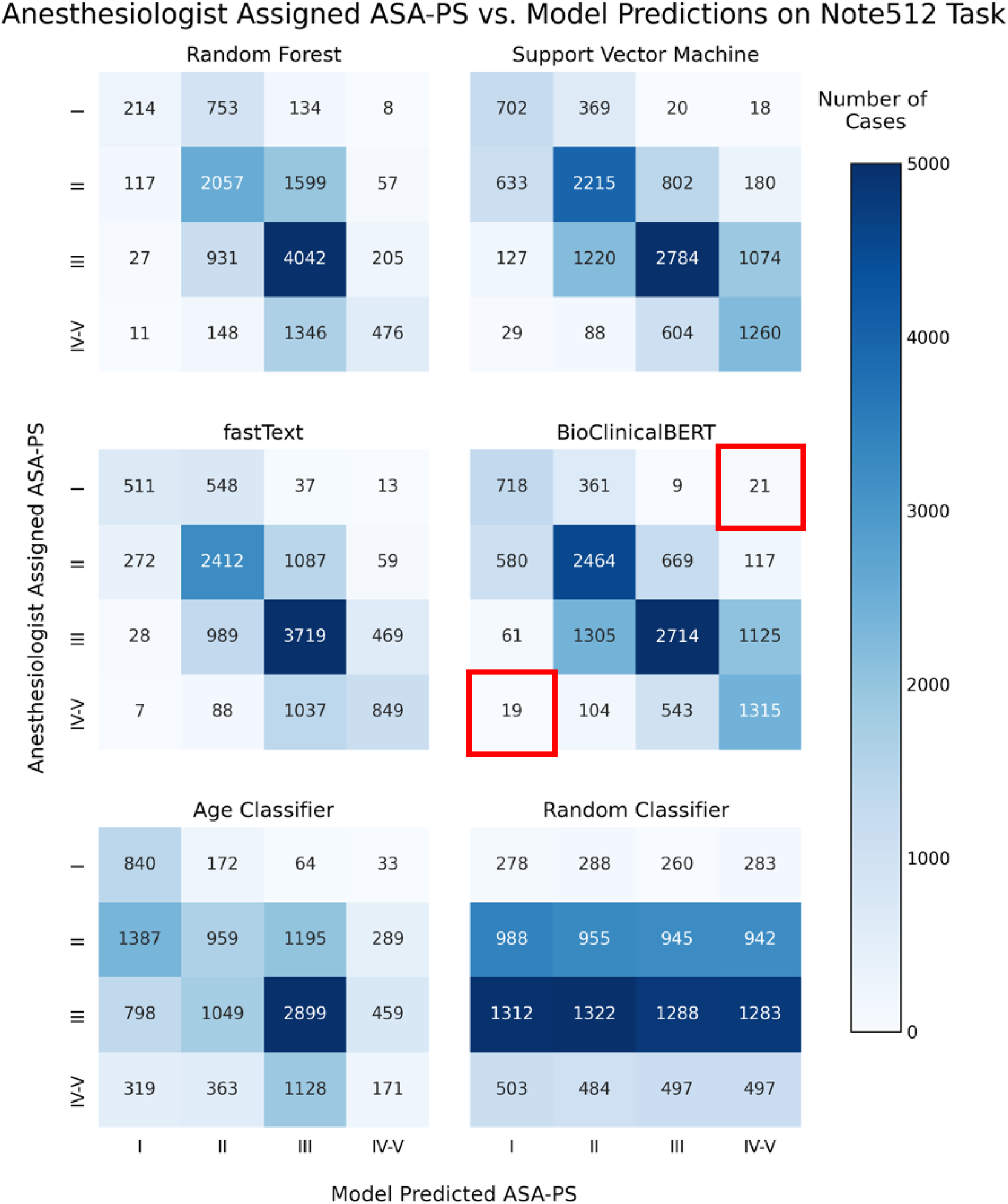
4-by-4 contingency tables for each model architecture on the Note512 task. The vertical axis corresponds to modified ASA-PS recorded in the anesthetic record by the anesthesiologist. The horizontal axis corresponds to the model predicted modified ASA-PS. Numbers in the table represent case count from the test set and show how these cases are distributed based on model prediction and actual ASA-PS recorded in the anesthetic record. Cells outlined in red in the BioClinicalBERT contingency table correspond to our definition of catastrophic errors. The 21 cases where anesthesiologist assigned ASA I and BioClinicalBERT model predicted ASA IV-V comprise 1.7% of cases. The 19 cases where anesthesiologist assigned ASA IV-V and BioClinicalBERT model predicted ASA I comprise 1.6% of cases.

**Figure 2:**
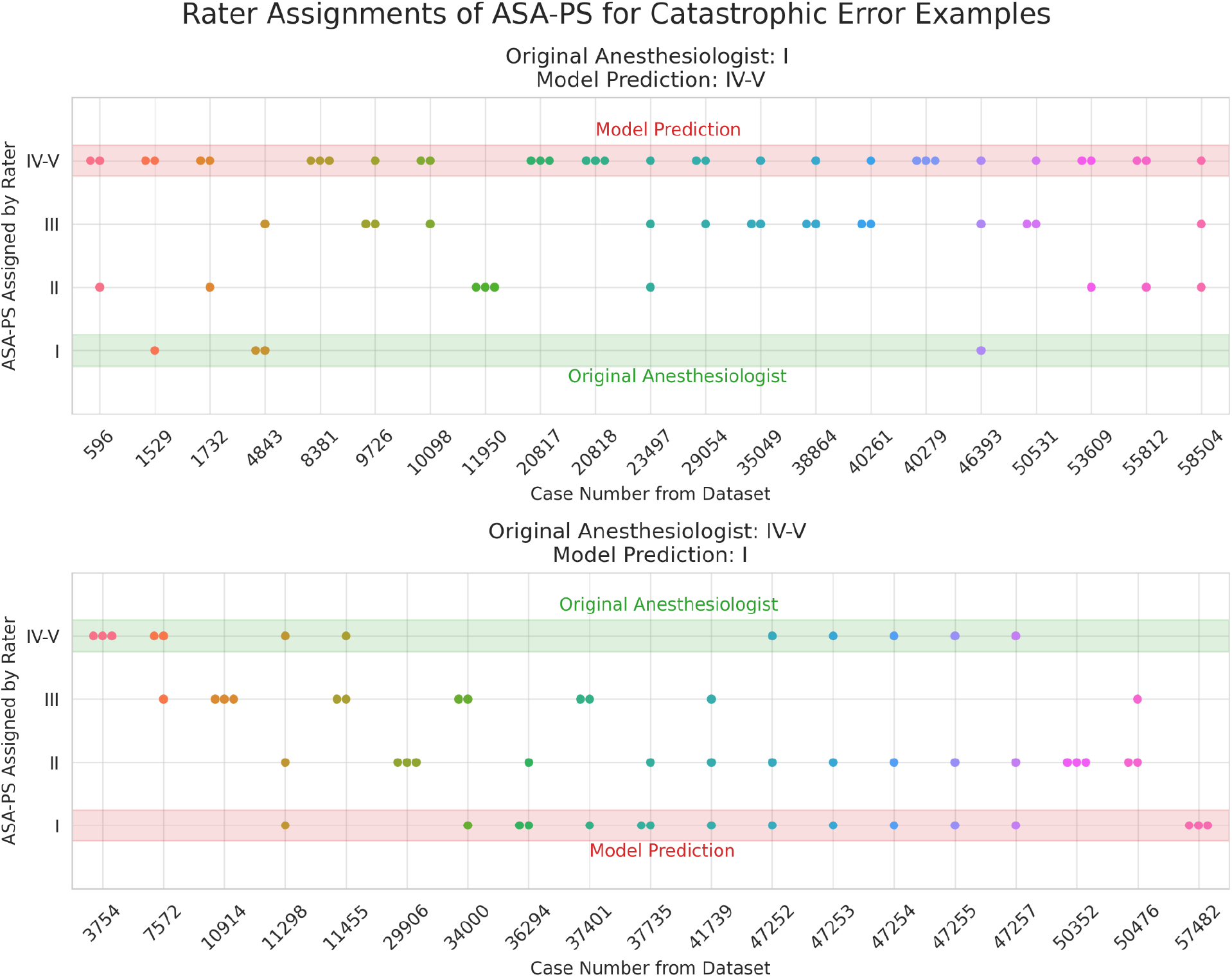
Rater assignments of ASA-PS for catastrophic error examples from the BioClinicalBERT model on Note512 task. Top plot shows scenario where model prediction is ASA IV-V, but original anesthesiologist assigned case ASA I. Bottom plot shows scenario where model prediction is ASA I, but original anesthesiologist assigned case ASA IV-V. Three anesthesiologist raters were asked to read the input text from the Note512 task and assign an ASA-PS for each of the catastrophic error examples. For each case, a dot marks a rater’s ASA-PS assignment. The model’s prediction and original anesthesiologist ASA-PS is shown as a highlighted region overlaid on the plots. Shapley feature attribution visualizations are shown for cases #57482 (Figure 3, eFigure 6), #41739 (eFigure 7), #11950 (eFigure 8), #29054 (eFigure 9).

Shapley values in Figure 3 provide clinically plausible explanations for model explanations, highlighting the directional probability of how specific input text contributes to predicting a specific ASA-PS. These feature attributions often provide clinically plausible explanations for why a model is making a wrong prediction and allows the clinician to evaluate the evidence the model is considering. Additional examples shown in eFigure 6, eFigure 7, eFigure 8, eFigure 9.

**Figure 3:**
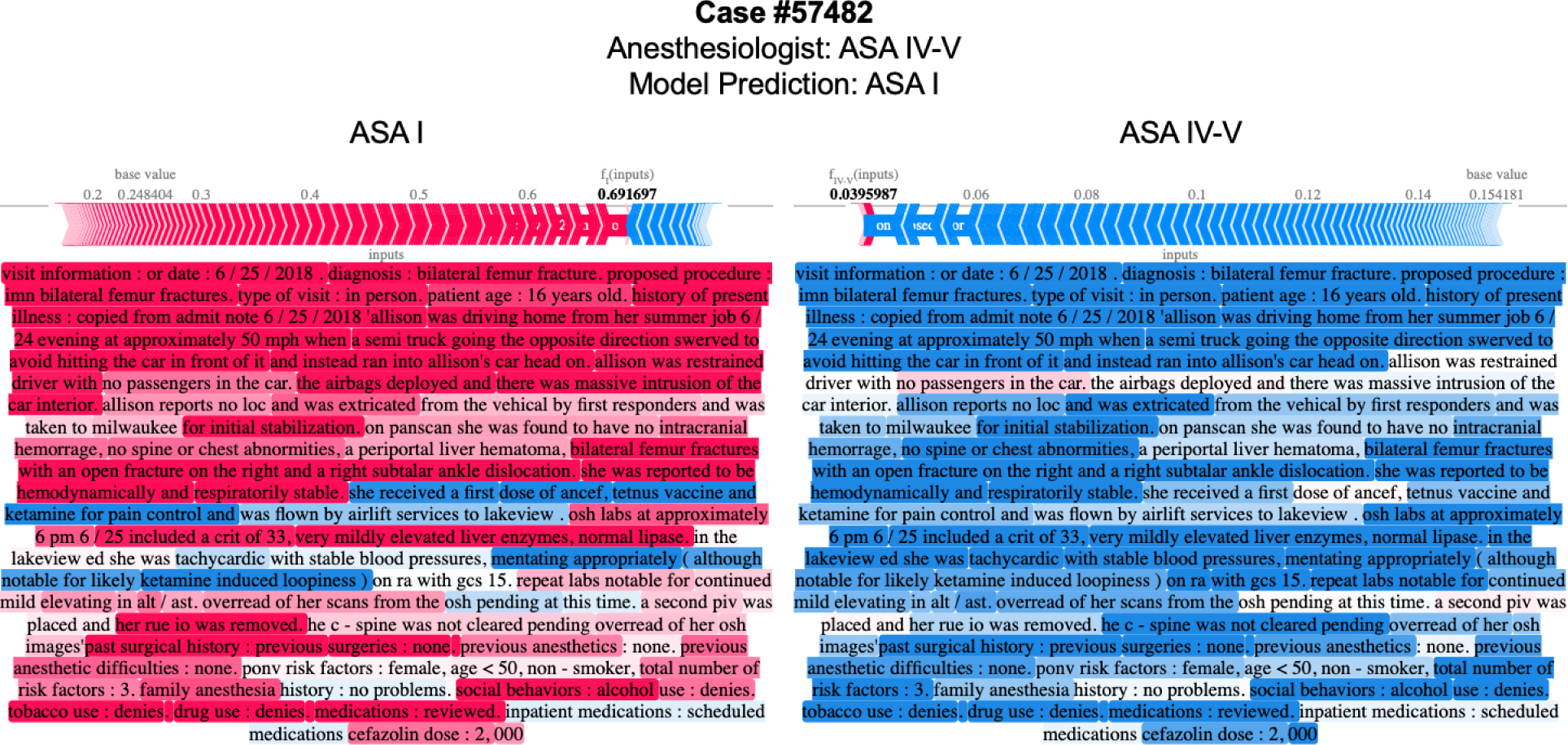
Attribution of input text features to predicting modified ASA-PS for the BioClinicalBERT model on Note512 task. Shapley values for each text token is shown to compare feature attributions to ASA I (top) and feature attributions to ASA IV-V (bottom). Red tokens positively support predicting the target ASA-PS whereas blue tokens do not support predicting the target ASA-PS. The magnitude and direction of support is overlaid on a force plot above the text. The baseline probability of predicting each class in the test set is shown as the “base value” on the force plot. The base value + sum of Shapley values from each token corresponds to the probability of predicting the ASA-PS and is shown as the bolded number. For simplicity, feature attributions to ASA II and III are omitted in this figure, but a full-visualization with all outcome ASA-PS for this text snippet is available in eFigure 6. Text examples are de-identified by replacing ages, dates, names, locations, and entities with pseudonyms to achieve data obfuscation while preserving structural similarity to the original passage.

## Discussion

Text classification techniques have undergone substantial evolution over the past decade. RF and SVM represent more rudimentary approaches that utilize bag-of-words and n-grams. These techniques are sensitive to word misspellings, cannot easily account for word order, have difficulty in capturing long-range references within sentences, and have difficulty in representing different meanings of a word when the same word appears in different contexts.^41–46^ Modern NLP techniques have overcome many of these challenges with: vector space representation of words^25,26,47–49^ and subword components^26,32,33,50^ as seen in the fastText model, attention mechanism^51,52^, and pretrained deep autoregressive neural networks^53–55^ such as transformer neural networks^56^. This has resulted in successful large language models such as BERT^34,57^ and the domain-specific BioClinicalBERT^35^.

Longer text length provides more information for the model to make an accurate prediction. Even though text snippets such as Diagnosis or Procedure may have high relevance for the illness severity of the patient, the better performance on longer input text sequences indicate that more information is generally better. This is similar to what is observed in the multifaceted practice of clinical medicine–where a patient’s overall clinical status is often better understood as the sum of many weaker but synergistic signals rather than a single descriptor. The limited input sequence length for BioClinicalBERT creates a performance ceiling as it limits the amount of information available to the model. Comparing Note and Note512 tasks, all other models that can utilize the full note have better performance when this input length is lifted with fastText being the top performer. These findings suggest that future development of a large language model similar to BioClinicalBERT capable of accepting a longer input context would likely have superior performance characteristics. fastText requires significantly less compute resources for model training and inference compared to BioClinicalBERT and remains a good option in lower resource settings. RF and SVM were our worst performing models, confirming that modern word vector and language model-based approaches are superior.

There is significant variability on the length and quality of clinical free-form text narrative written in the note, especially in the HPI section which is typically a clinician’s narrative of the patient’s medical status and need for the procedure. In some cases, the HPI section contains one or two words in length (eFigure 8), whereas in other cases it is a rich narrative (eFigure 6, eFigure 9). We believe that relatively poor performance in the ASA-PS prediction using HPI alone is a consequence of variability in documentation, as the model may have limited information for prediction if the note text does not richly capture the clinical scenario.

Models rarely make catastrophic errors. Erroneous predictions are typically adjacent to the ASA-PS assigned by the anesthesiologist, suggesting the model is making appropriate associations between freeform text predictors and the outcome variable (Figure 1). Examination of catastrophic errors from the BioClinicalBERT model on the Note512 task reveal that for both types of catastrophic errors–model predicts ASA I and original anesthesiologist assigned ASA IV-V, as well as the converse–we find that new anesthesiologist raters show greater concordance with the model predictions rather than the original anesthesiologist (Figure 2). Many of the catastrophic errors occurred with emergency cases. Shapley feature attributions for one of these catastrophic errors in Figure 3 reveal that in some cases the original anesthesiologist may have made the wrong assignment, or may have written a note that does not reflect the true clinical scenario. In this example, the original anesthesiologist assigned the case ASA IV-V, but the model predicted I. Feature attributions show the BioClinicalBERT model correctly identifies pertinent negatives on trauma exam, normal hematocrit of 33, and normal Glasgow Coma Scale (GCS) of 15 to all support a prediction for ASA I and against ASA IV-V. ^58^ In fact, all new anesthesiologist raters agree with the model rather than the original anesthesiologist. Examples like this suggest that the model performance may be underestimated by our evaluation metrics since our ground truth test set contains imperfect ASA-PS assignments. It illustrates how the model is robust against potentially faulty labels and has learned to make clinically appropriate ASA-PS predictions based on the input text

Shapley feature attributions reveal that the model is able to identify indirect indicators of a patient’s illness severity. For example, subcutaneous heparin is often administered for bed-bound inpatients to prevent the development of deep vein thrombosis. eFigure 8 depicts an example where the model learns to associate mention of subcutaneous heparin in the medication list with a higher ASA-PS, likely because hospitalized patients are generally more ill than outpatients who present to the hospital for same-day surgery. Similarly, the model learns the association between the broad spectrum antibiotic ertapenem with a higher ASA-PS as compared to narrow spectrum or prophylactic antibiotics such as metronidazole or cefazolin. These observations show that the model is able to identify and link these subtle indicators to a patient’s illness severity. Shapley value feature attributions prove to be an effective tool that enables clinicians to understand how a model makes its prediction from text predictors.

### Limitations

Our dataset is derived from a real-world EHR used to provide clinical care and includes human and computer generated errors. These issues include data entry and spelling, the use of abbreviations, references to other notes and test results not available to the model, and automatically generated/inserted text as part of a note template. The BioClinicalBERT model is limited to an input sequence of 512 tokens; future investigation is needed to understand if long-context large language models can achieve better performance. We also did not explore more advanced NLP models such as those that perform entity and relation extraction, which may further enhance the prediction performance. Finally, the ASA-PS is known to have only moderate interrater agreement among human anesthesiologists.^20,21^ Consequently, a perfect classification on this task is not possible since the ground truth labels derived from the EHR encapsulate this interrater variability. Further investigation is needed to explore the prediction of other outcome variables which may be less subject to interrater variability.

## Conclusions

NLP models can accurately predict a patient’s illness severity using only free-form text descriptions of patients without any manual data extraction. They can be automatically applied to entire panels of patients and serve as a perioperative risk stratification and clinical decision support tool to ensure patient safety. Illness severity predictions may also be used to reduce manual chart review overhead for medical billing. Shapley feature attributions produce explanations that logically support model predictions and are understandable to clinicians.

## Data Availability

Data will not be shared because the text dataset derived from electronic health records comprises personal identifiable information (PII) and protected health information (PHI). Code for experiments and results is publicly available.

https://github.com/philipchung/nlp-asa-prediction

## Data Availability

https://github.com/philipchung/nlp-asa-prediction

## Other Information

### Data Access, Responsibility, and Analysis

Philip Chung had full access to all the data in the study and takes responsibility for the integrity of the data and accuracy of the data analysis.

### Data Sharing Statement

Data will not be shared because the text dataset derived from electronic health records comprises personal identifiable information (PII) and protected health information (PHI). Code for experiments and results is publicly available at https://github.com/philipchung/nlp-asa-prediction.

## Acknowledgement Section

The authors would like to acknowledge: University of Washington Anesthesia Department’s Perioperative & Pain initiatives in Quality Safety Outcome group for assistance on data extraction and initial compute resources for data exploration, University of Washington Department of Medicine for computational environment support, Roland Lai and Robert Fabiano from University of Washington Research IT for creating a digital research environment within the Microsoft Azure Cloud where model development and experiments were performed, and the University of Washington Biomedical Natural Language Processing group for providing early feedback on experimental design and results.

## Author Contributions

Philip Chung: conception, experimental design, data acquisition, model development, data analysis, data interpretation

Christine Fong: data acquisition, data interpretation

Andrew Walters: data acquisition, data interpretation

Meliha Yetisgen: experimental design, data analysis, data interpretation

Vikas O’Reilly-Shah: experimental design, data analysis, data interpretation

## Supplemental Methods

Details on the approach taken for each of the four model architectures.

### Random Forest

Text input was preprocessed into a unigram and bigram count matrix followed by TFIDF transform.^23^ Random forest classifier from the Scikit-learn^59^ python library was used with minimization of gini impurity objective function and weighting each outcome class by inverse frequency to adjust for class imbalance. Hyperparameters tuned include: number of trees and number of features when looking for best split. Defaults were used for all other model parameters.

### Support Vector Machine

Text input was preprocessed into a unigram and bigram count matrix followed by TFIDF transform.^23^ LinearSVC^60^ from the Scikit-learn^59^ python library was used with minimization of squared hinge loss with L2 penalty^61^ and weighting each outcome class by inverse frequency to adjust for class imbalance. Crammer-Singer approach was used for the multiclass strategy.^62^ The “C” regularization strength parameter was tuned as a hyperparameter. Defaults were used for all other model parameters.

### fastText

Text was directly input into the fastText classification model, which internally combines word and sub-word vector representations using continuous bag-of-words^47^ and softmax with negative sampling loss^48^ objective function. Hyperparameters tuned include: learning rate, learning rate update rate, word vector dimension size, context window size, number of negatives sampled, number of epochs. Defaults were used for all other model parameters.

### BioClinicalBERT

Text was tokenized using WordPiece tokenizer^32–34^ and then used as input to a pre-trained BioClinicalBERT model^35^ with the addition of ASA-PS and Emergency modifier prediction heads, each consisting of a linear and softmax layer, for our specific ASA-PS prediction task (eFigure 3). These prediction heads were jointly optimized with AdamW optimizer^63^ during training using a weighted average of the cross-entropy loss from each prediction head; the weight of ASA-PS was held constant at 1.0 and the weight of the emergency modifier head was tuned as a hyperparameter. Cross-entropy loss is weighed by inverse class frequency to adjust for class imbalance. Both tokenizer and model are based on the Hugging Face^64^ python implementation with GPU acceleration enabled by PyTorch^65^ and PyTorch Lightning^66^. Tokenizer and model sequence length were set to the maximum of 512 tokens for the pretrained model. Longer input text sequences were truncated to this length. Hyperparameters tuned include: emergency head weight, batch size, learning rate, weight decay, dropout, gradient clipping, and number of epochs. ASHA^67^ with a reduction factor of 3 was used to tune up to 4 instances of the same model with different hyperparameters in parallel.

## Supplemental Figures

**eFigure 1:**
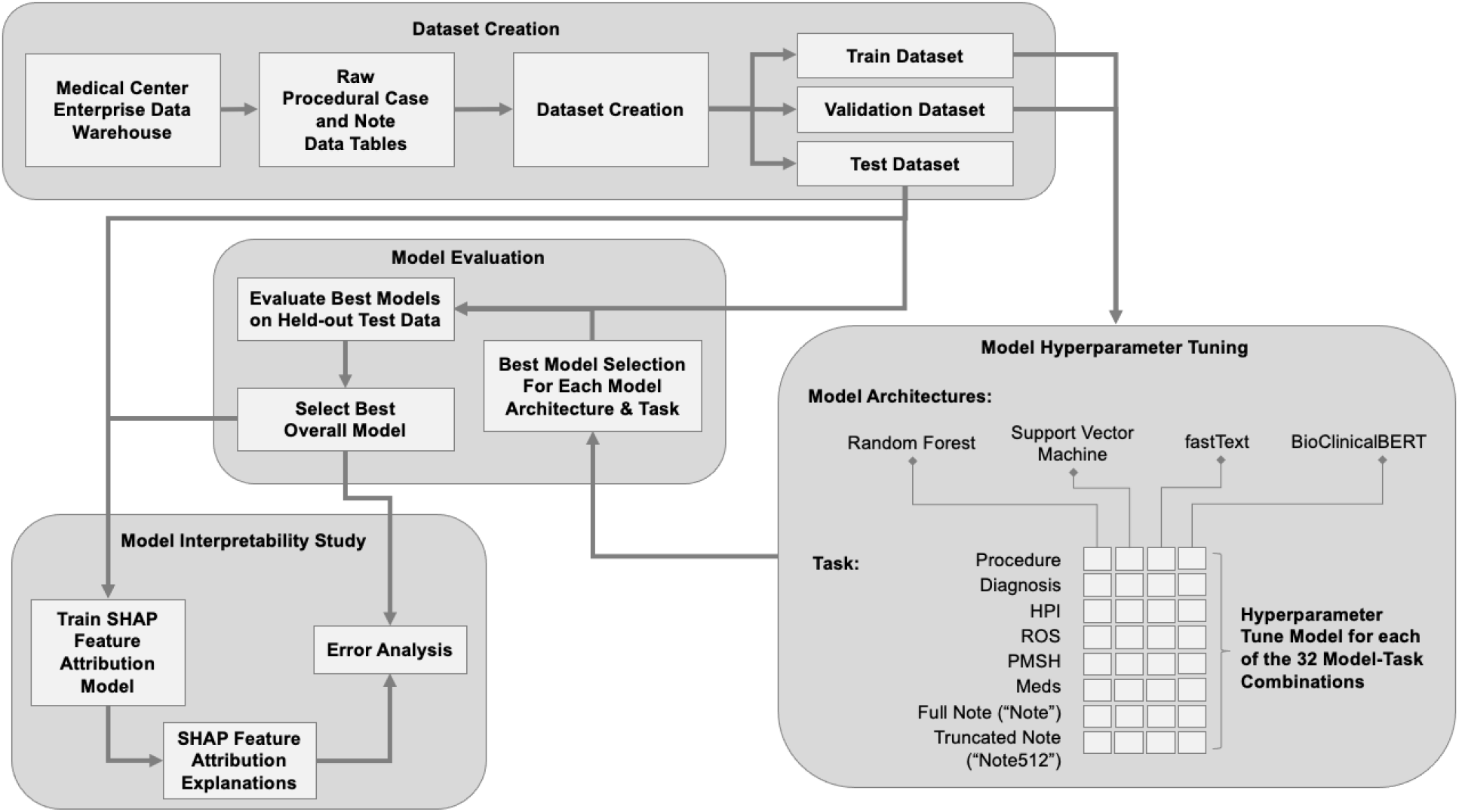
Flowchart of study design: dataset creation, model development, evaluation, and interpretation.

**eFigure 2:**
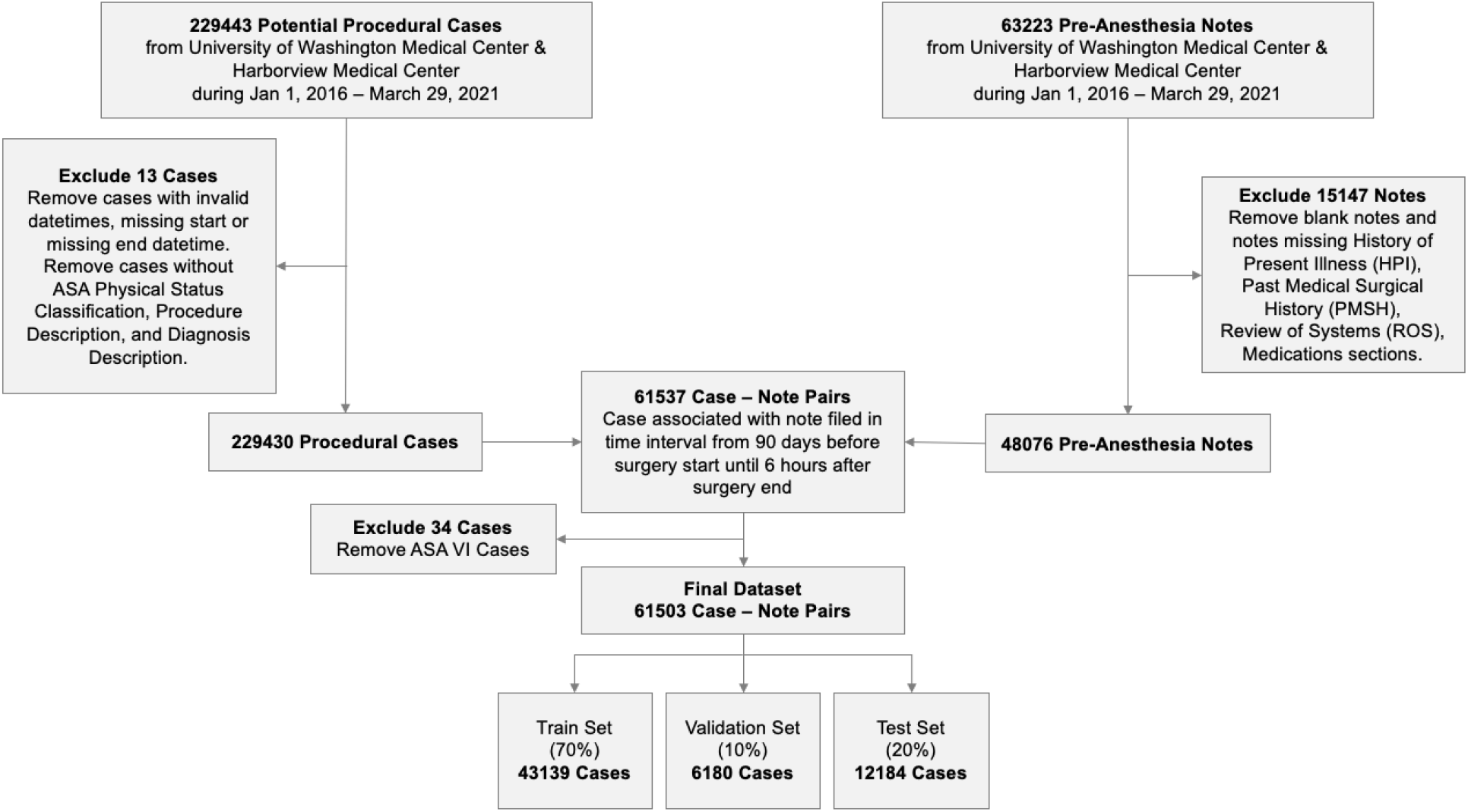
CONSORT Flow Diagram for Dataset Creation. If a patient has multiple procedural cases and pre-anesthesia notes, all of a patient’s cases and notes are allocated to the same data split.

**eFigure 3:**
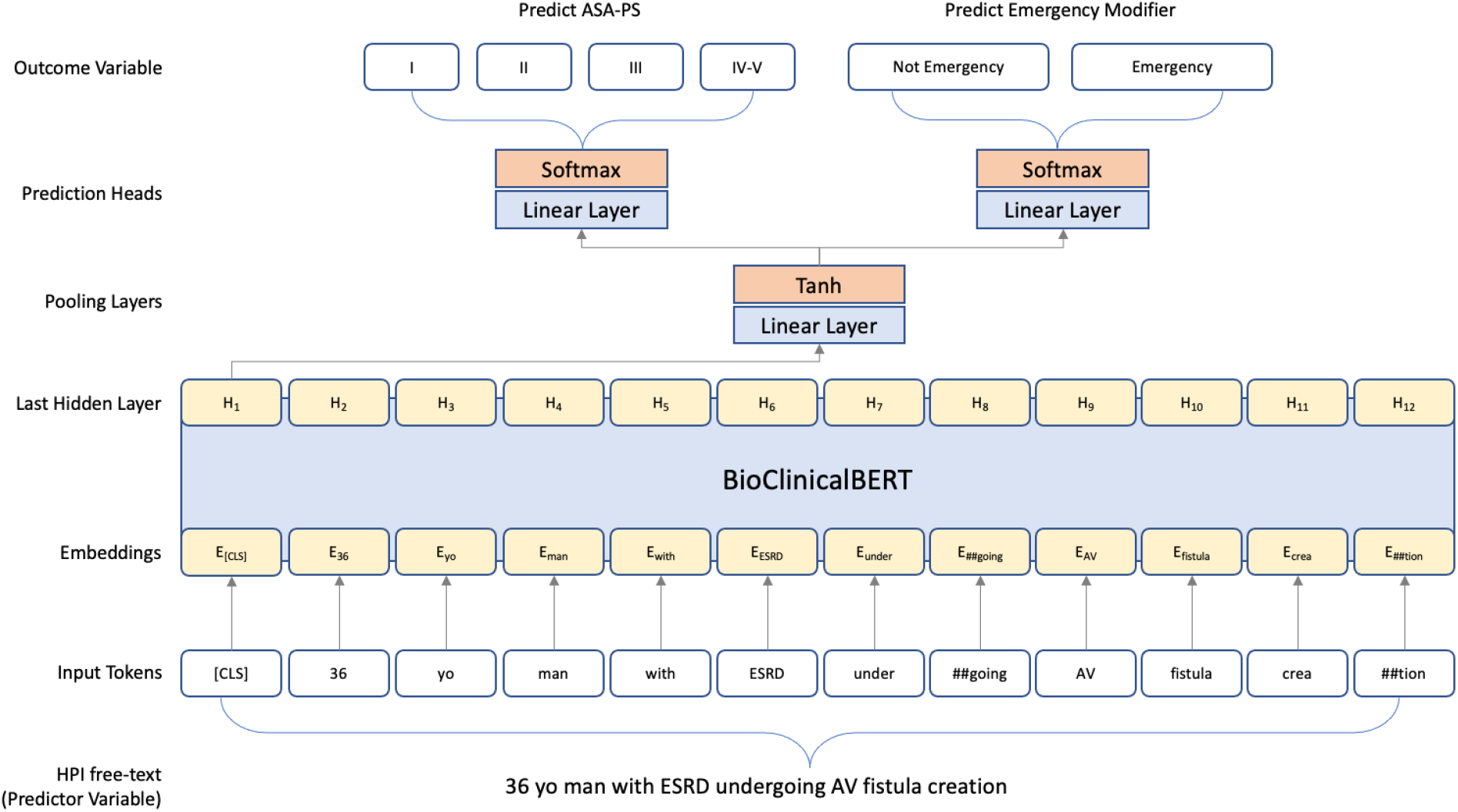
BioClinicalBERT Model Architecture with additional prediction heads for fine-tuning and prediction of modified ASA-PS

**eFigure 4:**
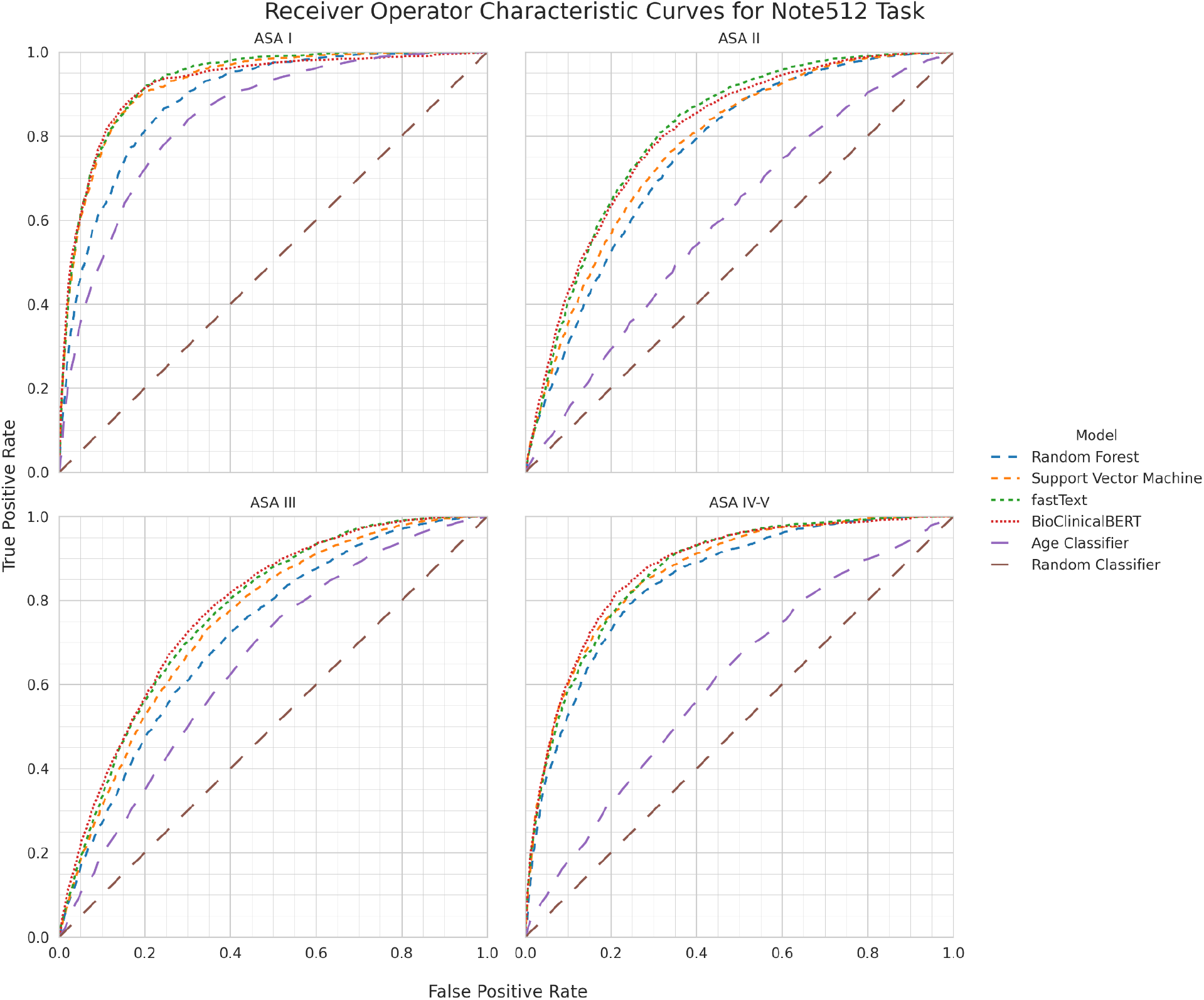
ROC performance of each model architecture on the Note512 task compared to baseline models. Each plot depicts model performance for predicting a specific ASA-PS.

**eFigure 5:**
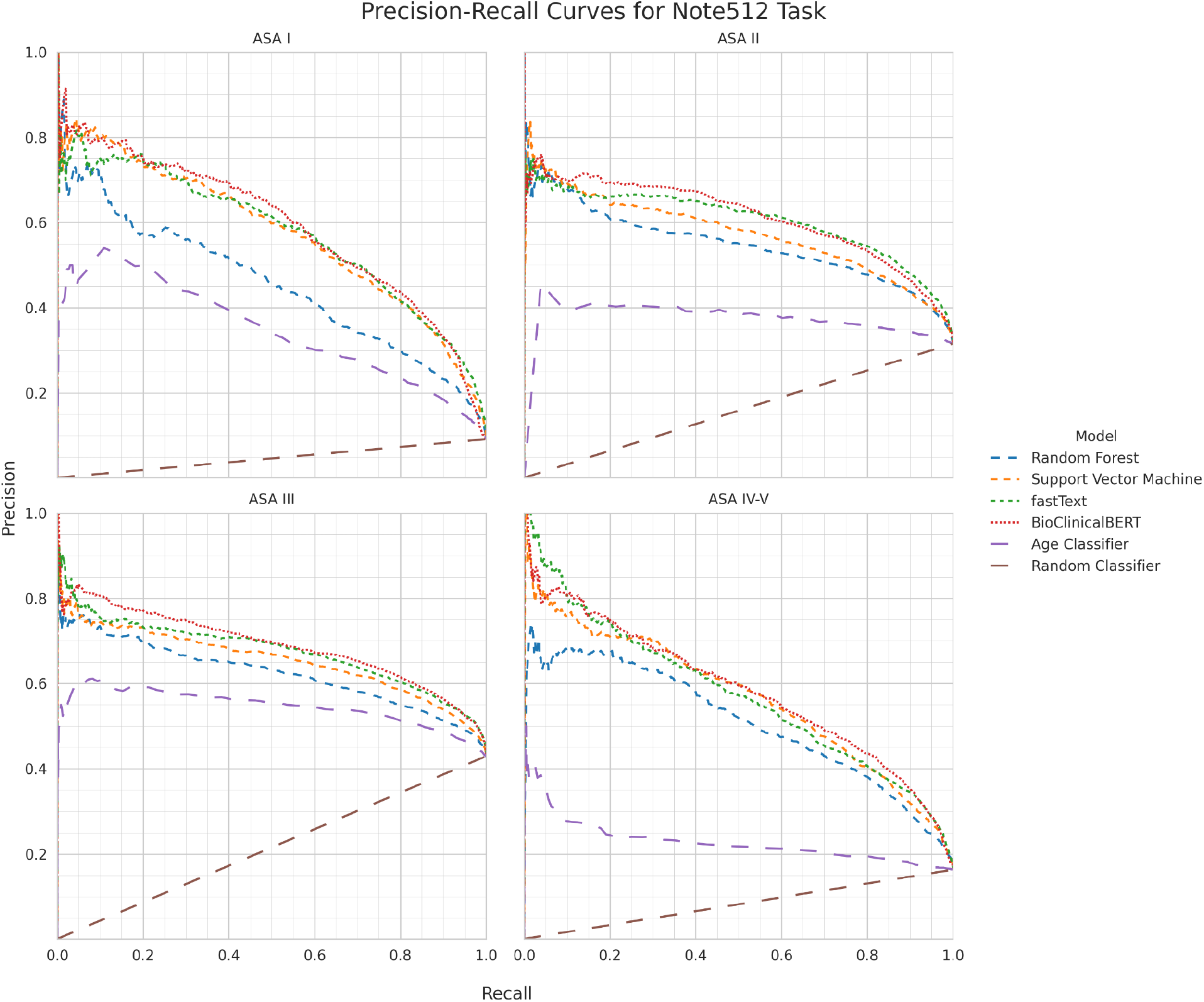
Precision-recall curve performance of each model architecture on the Note512 task compared to baseline models. Each plot depicts model performance for predicting a specific ASA-PS.

**eFigure 6:**
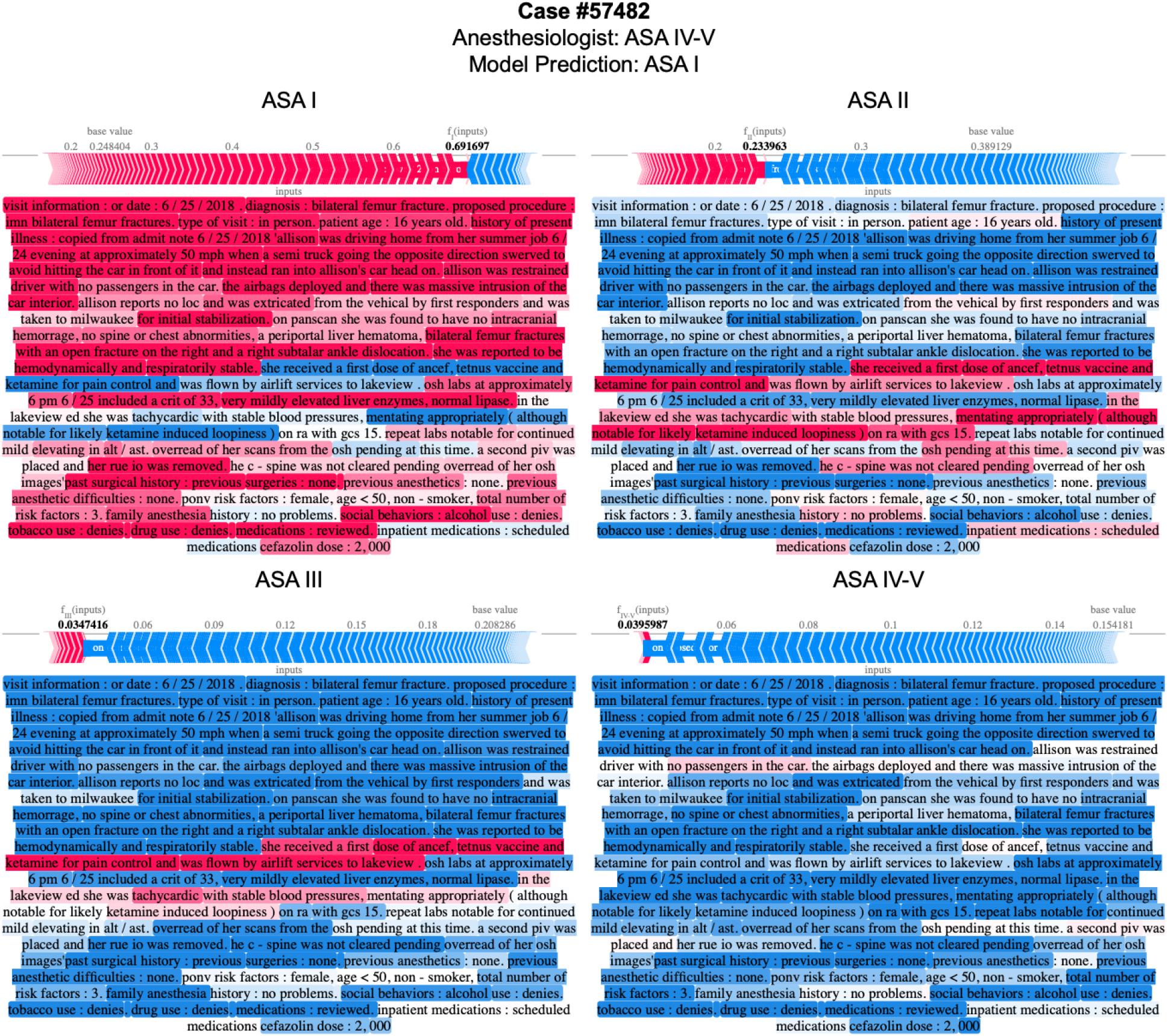
Attribution of input text features to predicting modified ASA-PS for the BioClinicalBERT model on Note512 task. Model prediction is ASA I, Anesthesiologist assigned case ASA IV-V. Notable findings include the model focusing on pertinent negatives on trauma exam and imaging findings and a normal hematocrit of 33 all of which support predicting a ASA-PS I. The same pertinent negatives as well as a Glasgow Coma Scale (GCS) of 15 are negatively Shapley values for ASA-PS IV-V, which reduce the probability of predicting ASA IV-V. Despite the anesthesiologist’s assignment of ASA IV-V, the text description does not suggest the patient has severe systemic disease with constant threat to life (ASA IV) or is moribund and requires the operation to survive (ASA V). Text examples are de-identified by replacing ages, dates, names, locations, and entities with pseudonyms to achieve data obfuscation while preserving structural similarity to the original passage.

**eFigure 7:**
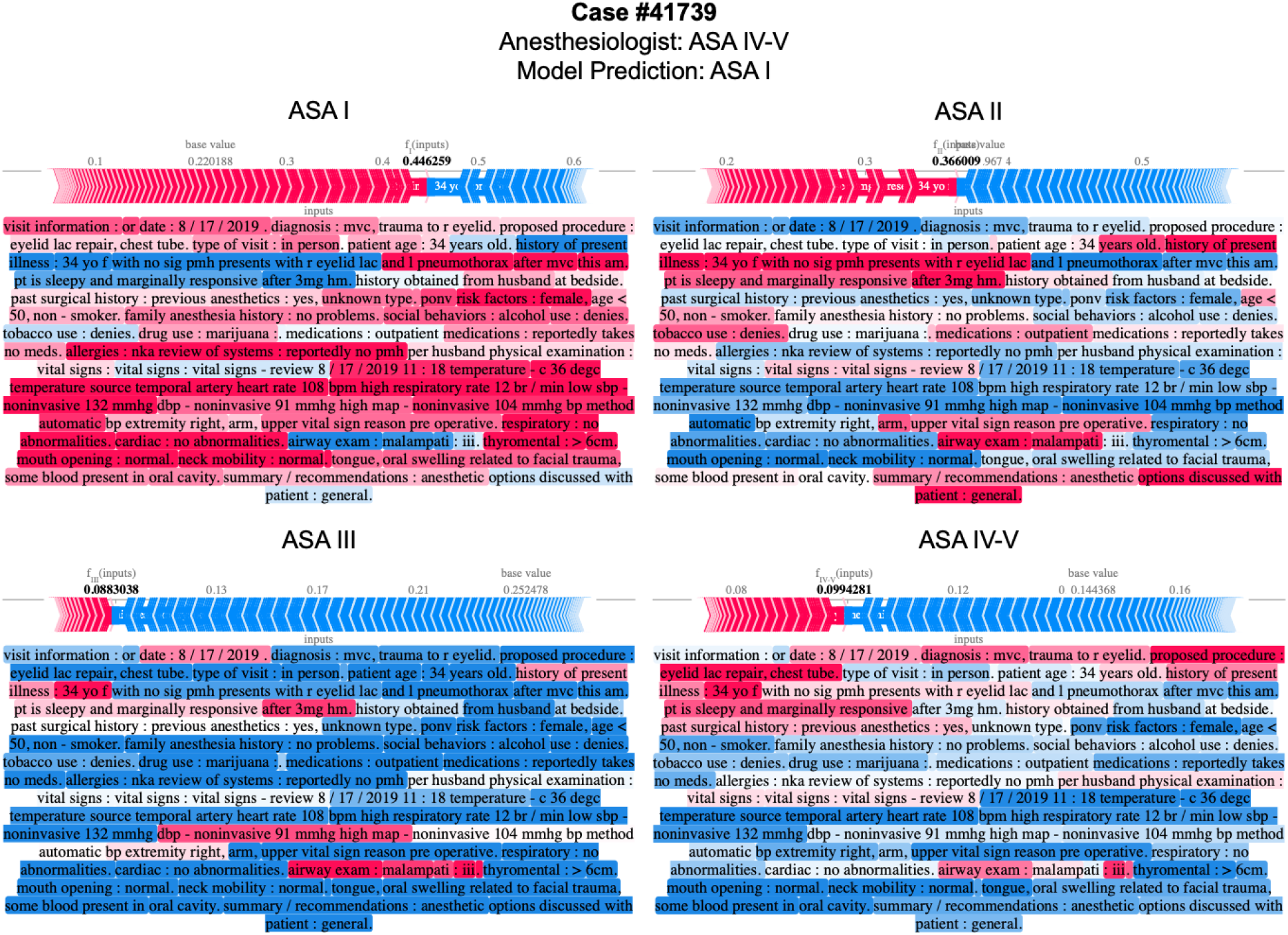
Attribution of input text features to predicting modified ASA-PS for the BioClinicalBERT model on Note512 task. Model prediction is ASA I, Anesthesiologist assigned case ASA IV-V. Notable findings include the model associating chest tube with ASA IV-V. The model has trouble with consistently attributing the multiple mentions of eyelid laceration with a specific ASA-PS. The model may be inappropriately assigning mention of left pneumothorax to ASAI. This example depicts a challenge for the model in which a relatively minor injury (eyelid laceration) is simultaneously present with a potentially severe injury (pneumothorax), though the severity of the pneumothorax is not mentioned and thus the text predominantly supports ASA I (healthy) or ASA II (mild systemic disease). This kind of mixed illness/injury example coupled with a narrative that does not clearly describe disease severity may be a struggle for the model. Text examples are de-identified by replacing ages, dates, names, locations, and entities with pseudonyms to achieve data obfuscation while preserving structural similarity to the original passage.

**eFigure 8:**
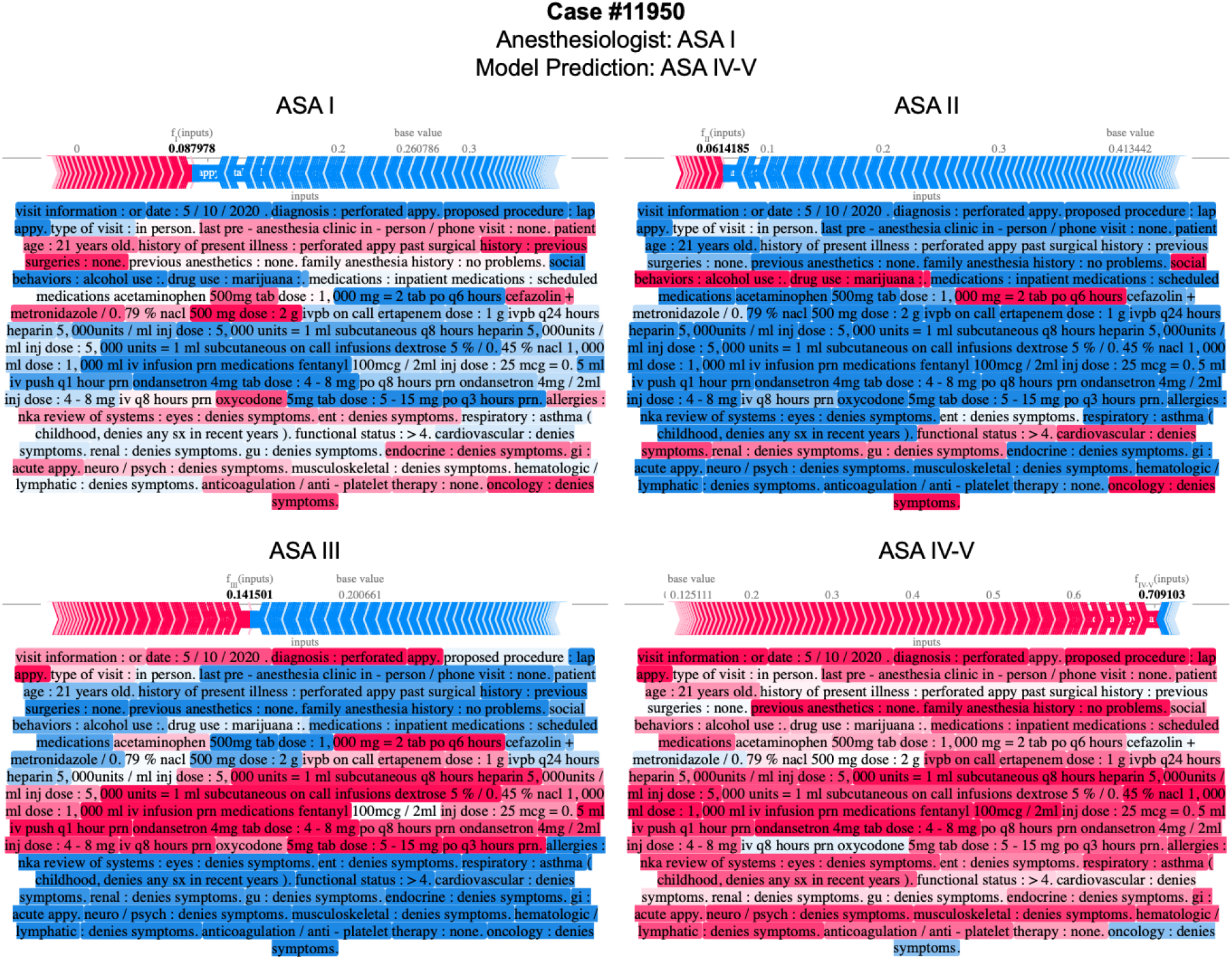
Attribution of input text features to predicting modified ASA-PS for the BioClinicalBERT model on Note512 task. Model prediction is ASA IV-V, Anesthesiologist assigned case ASA I. Notable findings include: young age associated with ASA I and ASA IV-V, but negatively associated with ASA II and III; diagnosis of perforated appendix and procedure of laparoscopic appendectomy negatively associated with ASA I and positively associated with higher ASA-PS; model identifying broad-spectrum antibiotics such as ertapenem to be associated with ASA IV-V, but narrower-spectrum antibiotics such as metronidazole, cefazolin to be heavily associated with ASA I; inpatient medications such as subcutaneous heparin and ondansetron negatively associated with lower ASA-PS and positively associated with higher ASA-PS. Text examples are de-identified by replacing ages, dates, names, locations, and entities with pseudonyms to achieve data obfuscation while preserving structural similarity to the original passage.

**eFigure 9:**
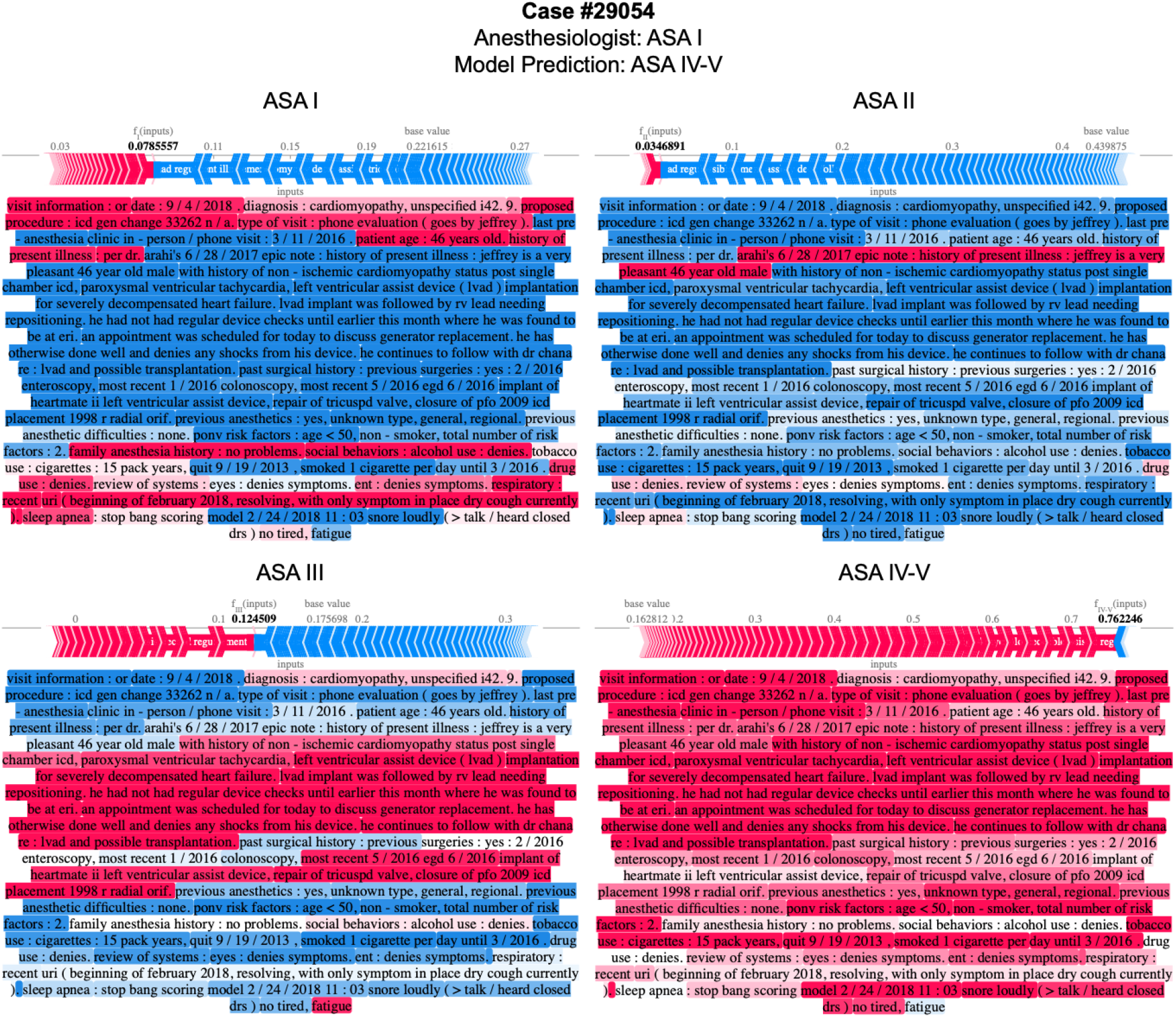
Attribution of input text features to predicting modified ASA-PS for the BioClinicalBERT model on Note512 task. Model prediction is ASA IV-V, Anesthesiologist assigned case ASA I. Notable findings include medical conditions and interventions associated with higher ASA-PS such as cardiomyopathy, internal cardiac defibrillator (ICD) generator change, paroxysmal ventricular tachycardia, left ventricular assist device (LVAD), heart failure, possible transplantation, tricuspid valve repair, and patent foramen ovale (PFO) closure; history of chronic cigarette smoking and snoring associated with ASA IV-V. The text description is at least ASA III (severe systemic illness), and can be argued to be ASA IV (severe systemic disease with constant threat to life) if heart failure is progressively worsening. In this example the model appears to make a more appropriate ASA-PS prediction than the anesthesiologist. Text examples are de-identified by replacing ages, dates, names, locations, and entities with pseudonyms to achieve data obfuscation while preserving structural similarity to the original passage.

**eTable 1:**
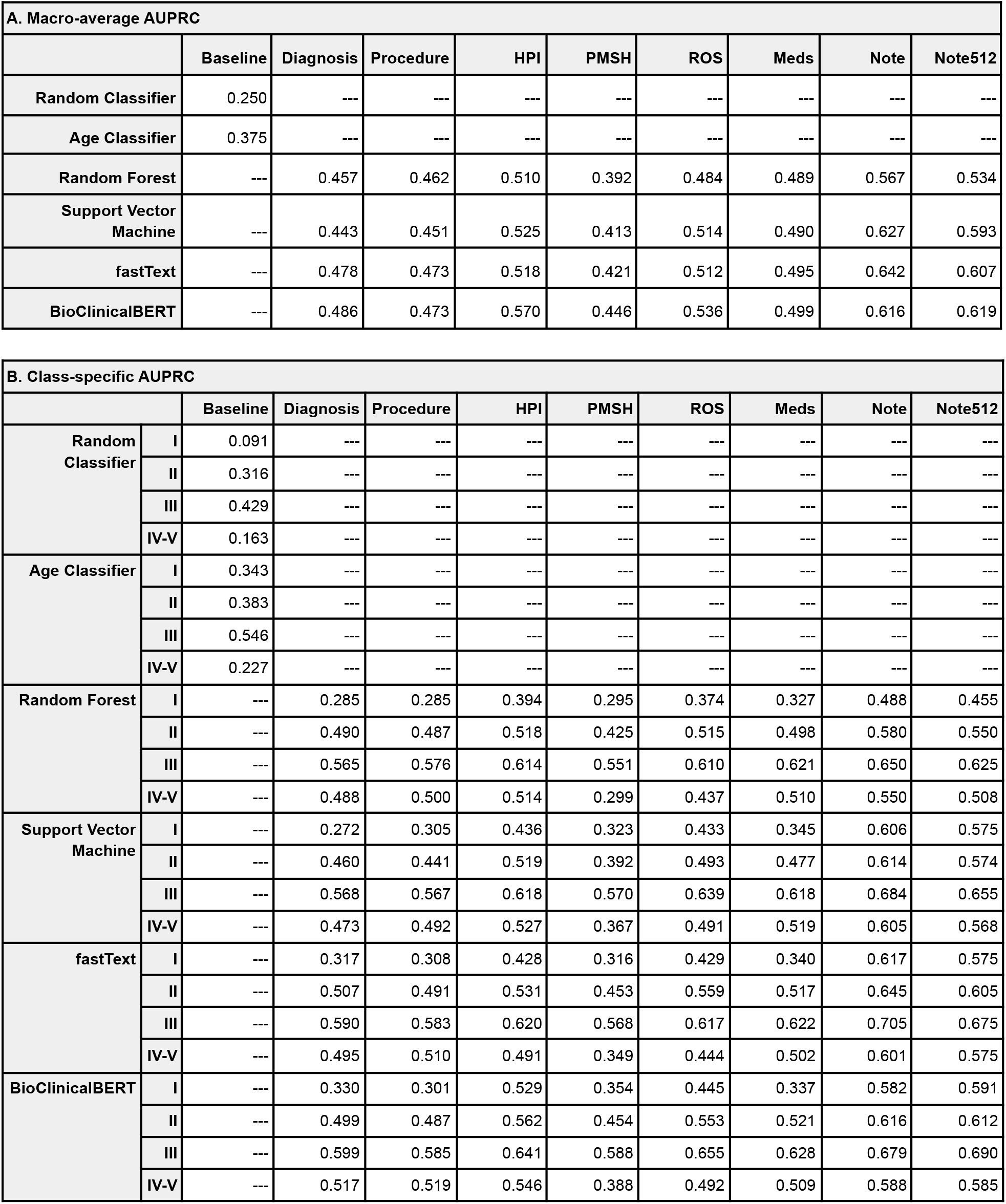
(A) Macro-average AUPRC and (B) class-specific AUPRC for each model architecture and task on the held-out test set compared to baseline models. Random Classifier serves as a negative control baseline. Age classifier serves as a simple clinical baseline since ASA-PS typically increases as a patient ages and has increased medical comorbidities.

**eTable 2:**
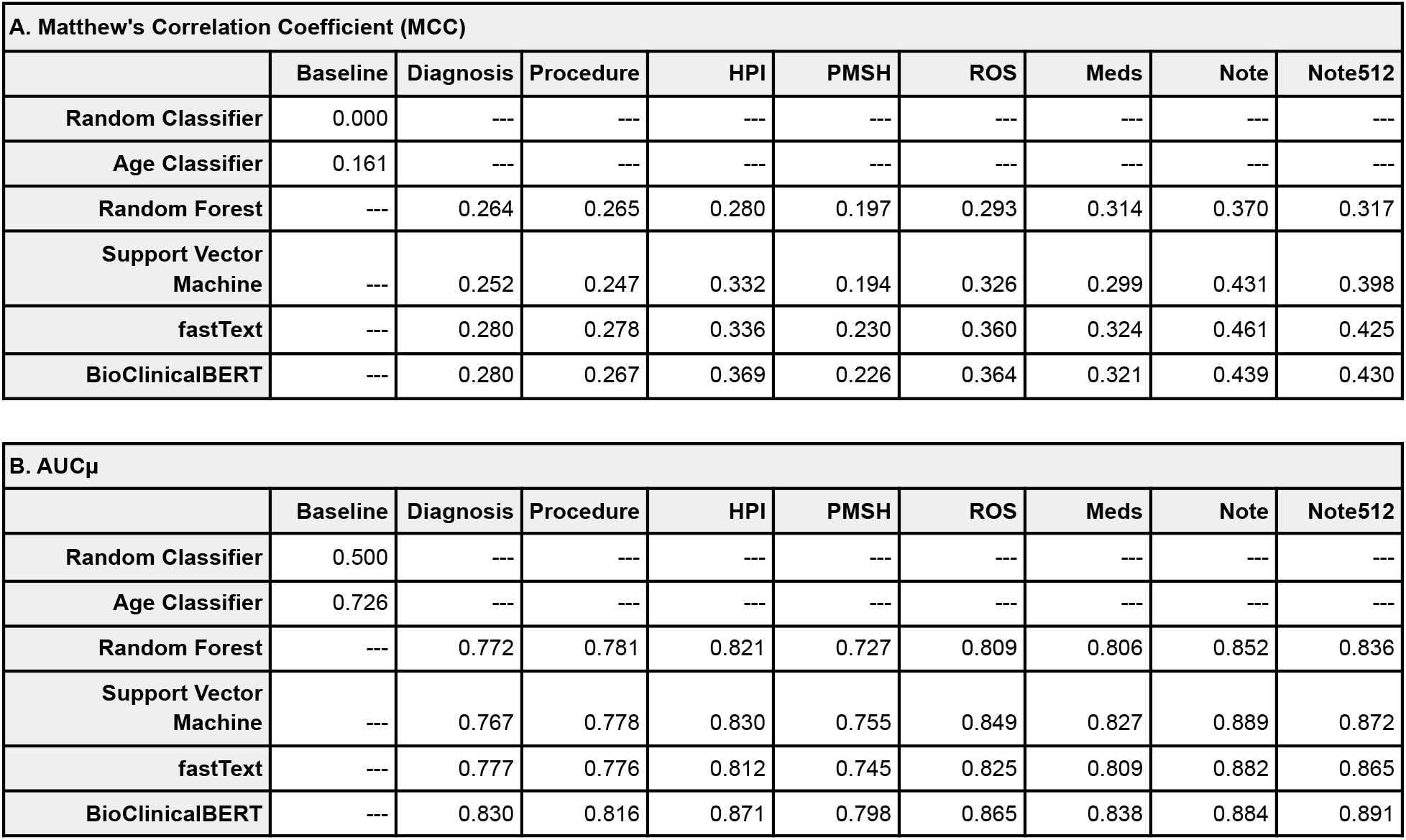
(A) Matthew’s correlation coefficient (MCC) and (B) AUCμ for each model architecture and task on the held-out test set compared to baseline models. MCC is a categorical analog of Pearson’s correlation coefficient. AUCμ is a multiclass generalization of AUROC and U statistic and is more theoretically grounded than macro-average AUROC, but less commonly reported.

**eTable 3:**
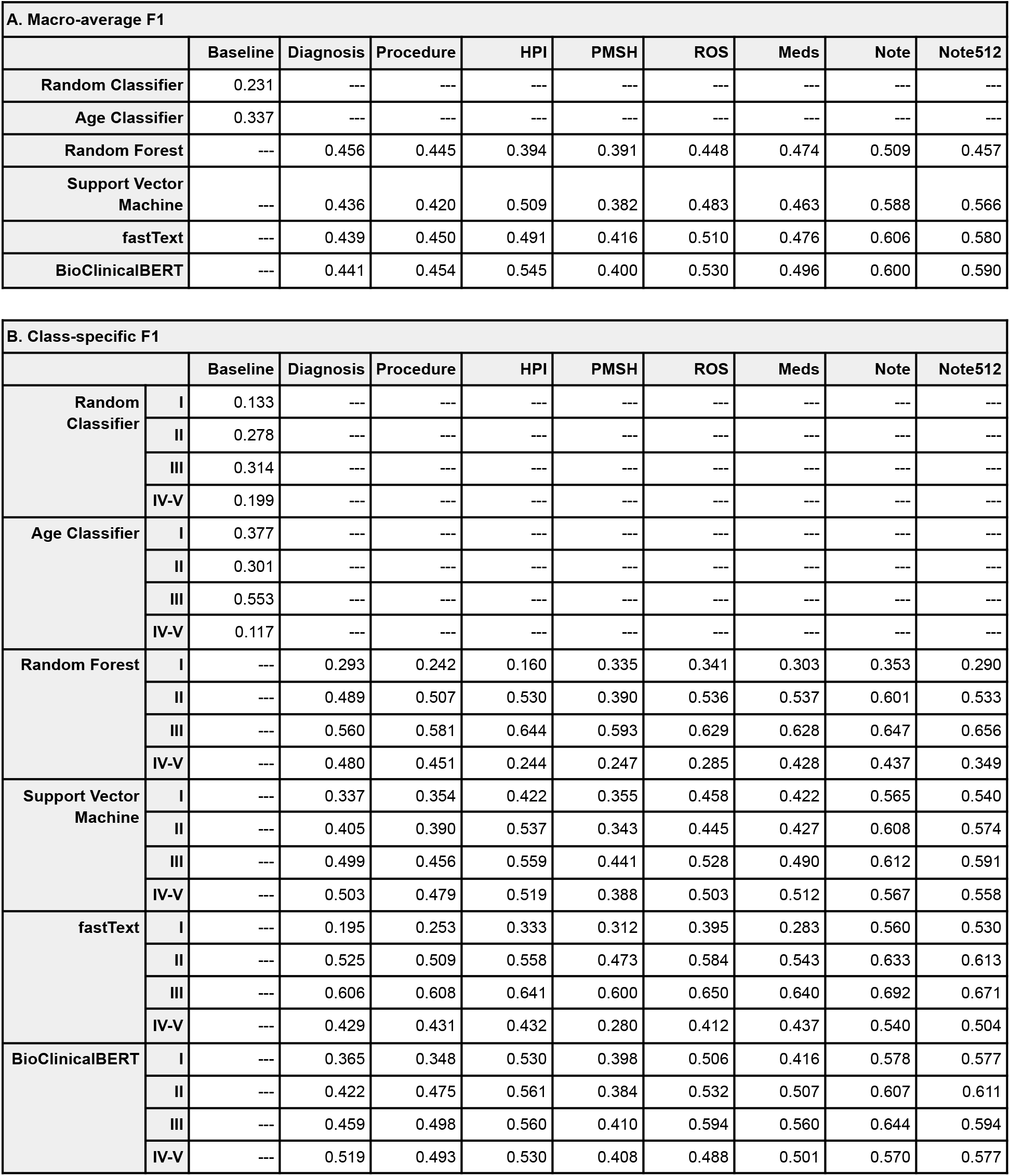
(A) Macro-average F1 and (B) class-specific F1 for each model architecture and task on the held-out test set compared to baseline models.

**eTable 4:**
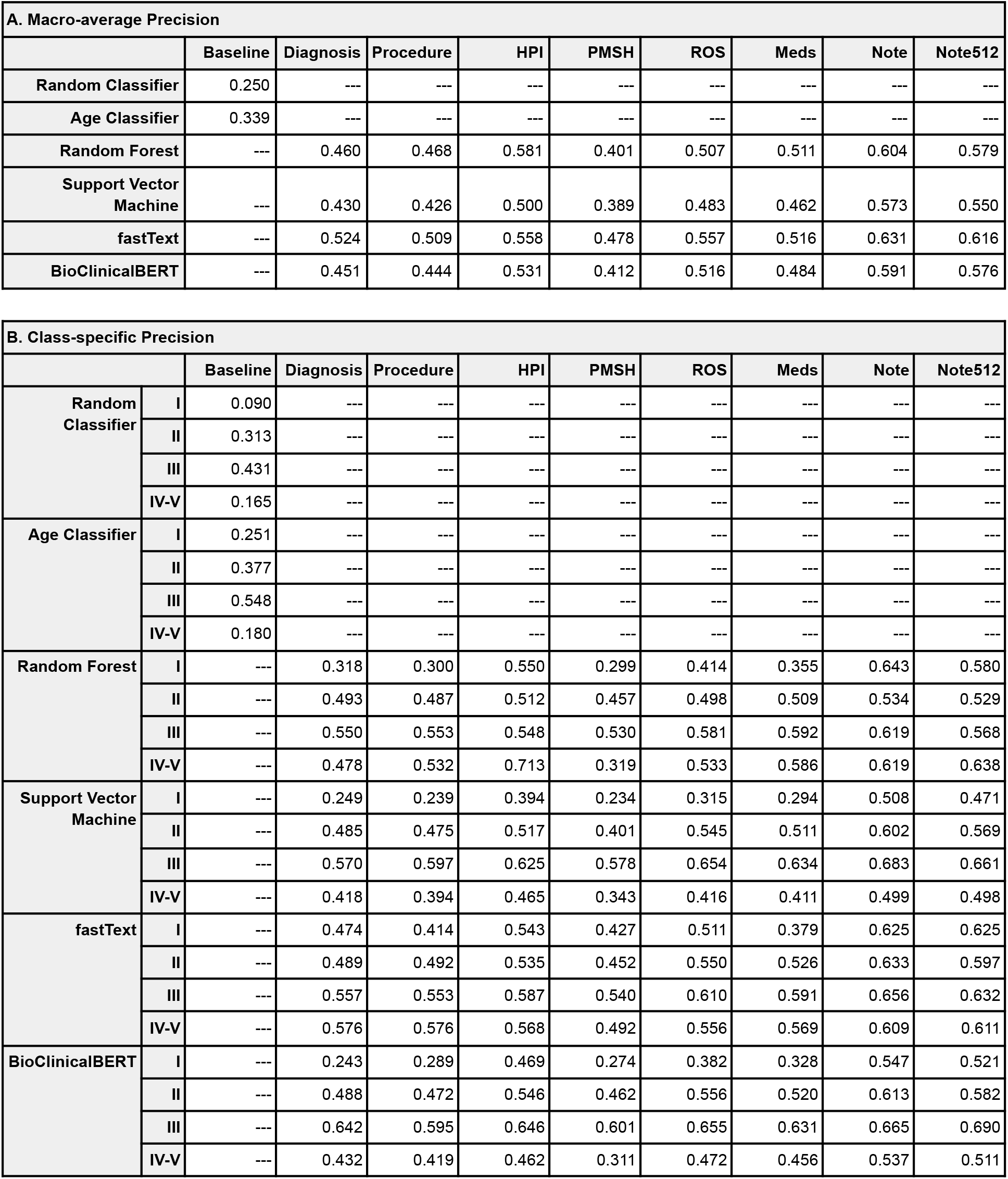
(A) Macro-average precision and (B) class-specific precision for each model architecture and task on the held-out test set compared to baseline models.

**eTable 5:**
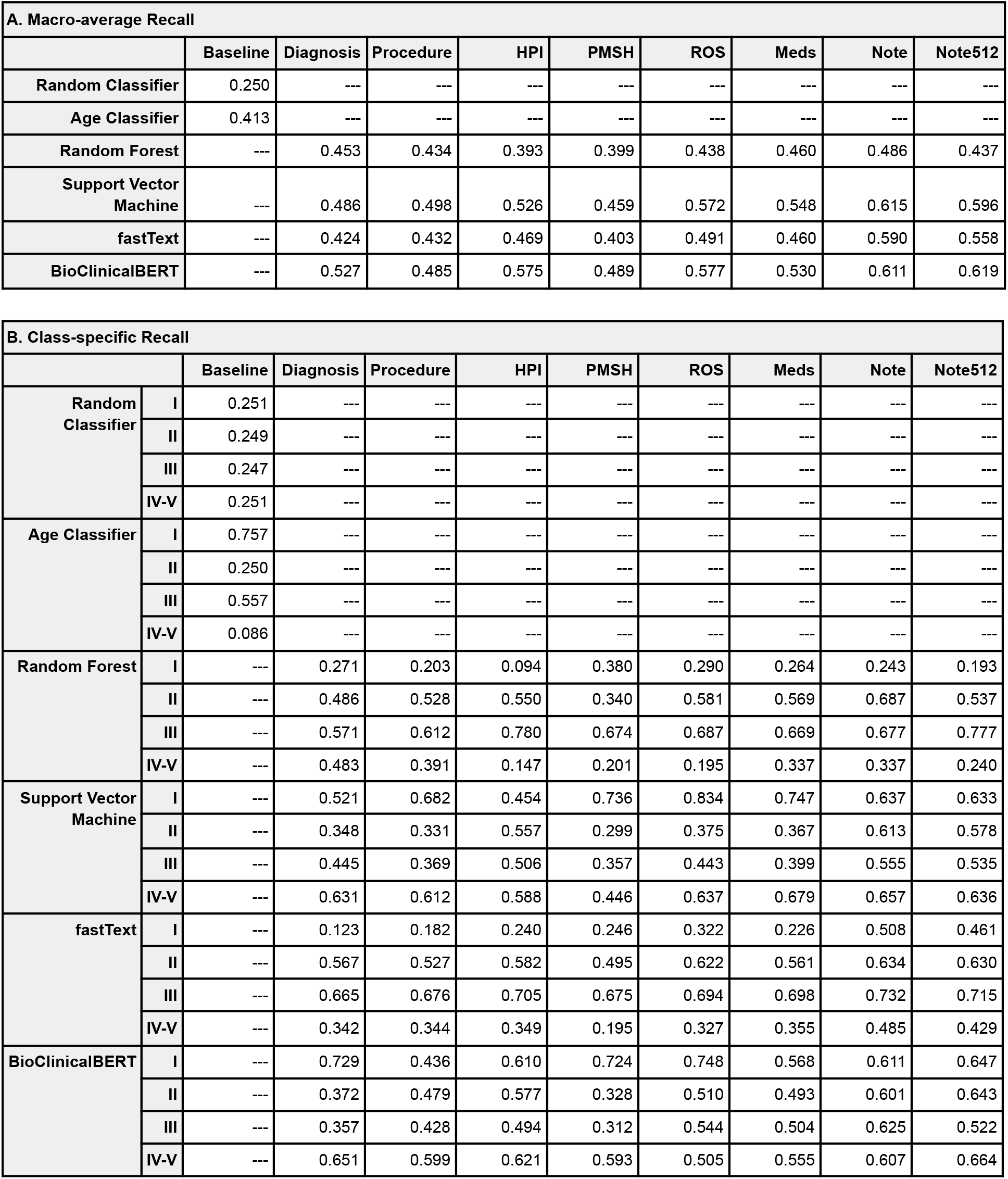
(A) Macro-average recall and (B) class-specific recall for each model architecture and task on the held-out test set compared to baseline models.

